# Detection and Characterization of Walking Bouts Using a Single Wrist-Worn Accelerometer in Free-living Conditions

**DOI:** 10.1101/2023.08.01.23293509

**Authors:** Andreas Brink-Kjær, Sajila D. Wickramaratne, Ankit Parekh, Emmanuel H. During

## Abstract

Detection and characterization of abnormalities of movement are important to develop a method for detecting early signs of Parkinson’s disease (PD). Most of the current research in detection of characteristic reduction of movements due to PD, known as parkinsonism, requires using a set of invasive sensors in a clinical or controlled environment. Actigraphy has been widely used in medical research as a non-invasive data acquisition method in free-living conditions for long periods of time. The proposed algorithm uses triaxial accelerometer data obtained through actigraphy to detect walking bouts at least 10 seconds long and characterize them using cadence and arm swing. Accurate detection of walking periods is the first step toward the characterization of movement based on gait abnormalities. The algorithm was based on a Walking Score (WS) derived using the value of the auto-correlation function (ACF) for the Resultant acceleration vector. The algorithm achieved a precision of 0.90, recall of 0.77, and F1 score of 0.83 compared to the expert scoring for walking bout detection. We additionally described a method to measure arm swing amplitude.

## I. Introduction

Rapid-eye-movement sleep behavior disorder (RBD) affects 1% of adults and is in most cases the early manifestation of a neurodegenerative disease, Parkinson’s disease (PD) or dementia with Lewy bodies [1][2]. Both conditions are characterized by the presence of “parkinsonism,” observed as a progressive restriction and slowing of movements and gait abnormalities, such as arm swing asymmetry, which conventionally requires physical examination by a trained neurologist. RBD causes frequent twitches during sleep, which, as shown in our prior work, can be accurately detected by wrist accelerometers [3]. Wrist-worn accelerometry could thus facilitate the diagnosis of patients with RBD in the general population, however a convenient and scalable method for detecting early signs of parkinsonism in this patient population is needed.

Several studies have used accelerometry for detecting parkinsonism, particularly differences in gait parameters in patients with PD or RBD against healthy controls. In the prodromal stage of isolated RBD, reduced cadence, increased stride time, swing time [4], step width variability, and step length asymmetry could be observed, in addition to differences in gait initiation [5], however these could be measured only in the clinical setting, using pressure sensor carpet. U-turn slowing is another PD characteristic, which only recently could be measured in free-living conditions [6], however this feature has only been tested with a hip-worn smartphone, and in RBD, only in the clinic setting under controlled conditions [7][8]. A study using a lower back sensor showed that higher step time variability and asymmetry of all gait characteristics can be detected years prior to PD diagnosis, but this also required standardized motor testing and is more invasive than a wrist sensor [9].

In comparison, very few studies have tested wrist accelerometry in PD or prodromal patients. Mirelman et al. found significant differences in arm swing in mutation carriers prior to any detectable parkinsonism but this was only noted under dual-task conditions and using gyroscopes [10]. The literature on wrist accelerometry in free-living conditions to measure motor changes associated with PD is scarce. One study reported reduced median arm movement power during non-gait activities [6], which may be interpreted as a reflection of bradykinesia. Another study using wrist accelerometer data from the United Kingdom (UK) Biobank and machine learning methods analyzing gait and low-movement data achieved good performance (area under the curve 0.85 when fused) for distinguishing PD from non-PD [11].

In summary, although most studies for measuring parkinsonism have been conducted in the in-clinic setting or have required controlled conditions and/or invasive sets of sensors, emerging data using a wristworn sensor suggests that analysis of gait and non-gait movements in free-living conditions is feasible, and could be used to monitor disease progression in RBD. This article presents an algorithm that uses single-wrist actigraphy to detect and characterize walking bouts in free-living conditions.

## II. Methods

The system overview is shown in Figure 1. The system consists of data acquisition, preprocessing, Walking Bout Detection, and Post Processing Phases. After the Post-processing phase, the detected walking bouts were used to calculate cadence and arm swing for the walking bouts. Each phase will be discussed in detail in the following sections.

**Figure 1.**
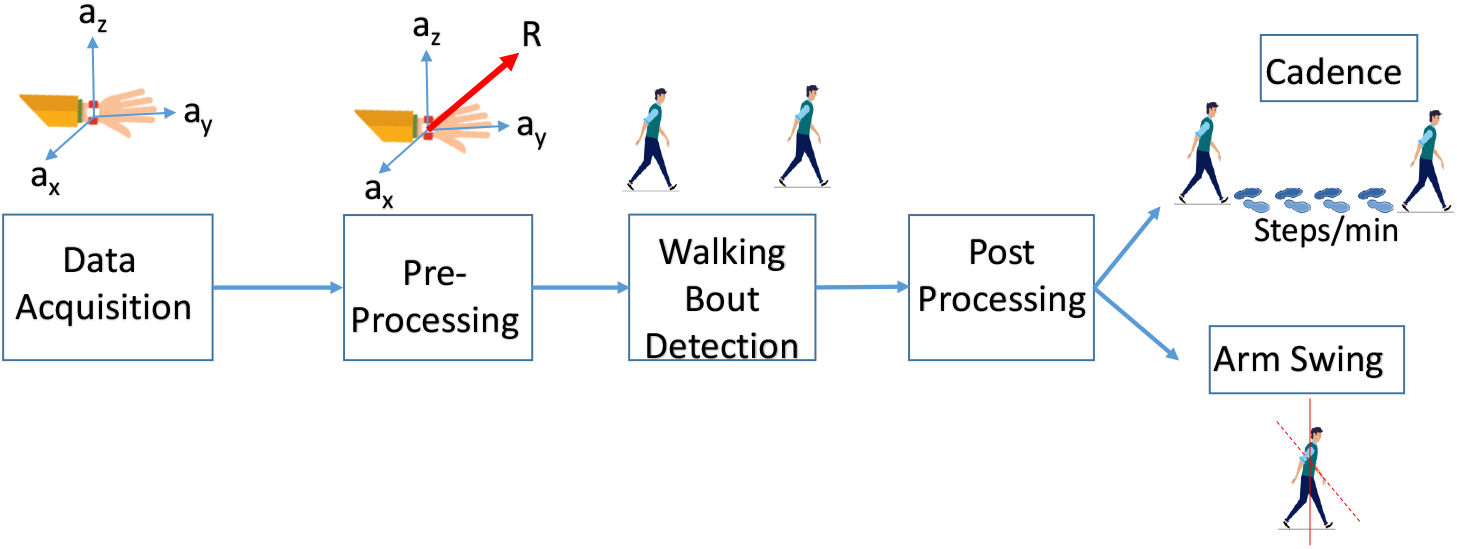
Overview of the proposed method for detection and characterization of walking bouts.

### II-A. Data Description

Eight healthy volunteer adult participants, 3 females and 5 males aged 26 to 46 (32.3 ± 7.2) years, were included in this study. Participants were instructed to wear a single AX-6 (Axivity, Ltd, Newcastle, UK) actigraphy device at the dominant wrist for at least 24 and up to 48 hours during weekdays and to follow their usual routines, and mark the start and end times of any continuous walking bout estimated to be longer than 1 minute.

Actigraphs recorded triaxial accelerations at a resolution of 25 Hz using a dynamic range of ±8 gravity (g). After all participants returned their device and the list of time stamps for their walking bouts, an independent reviewer visually inspected raw actigraphy data for each participant using the software Open Movement graphical user interface (OMGUI) and corrected or added any walking bout to ensure that all labels were accurate and, to the greatest extent possible, no obvious walking bout had been missed. Moreover, one of the 8 recordings was carefully reviewed and labelled in 10-second windows to capture any short meaningful walking bouts.

A total of 816.9 hours of actigraphy data was available for analysis, averaging 102.1 ±87.1 hours per participant. The average number of walking bouts was 4.3 *±* 2.5 per day and per participant, with durations ranging from 1 to 134 minutes and an average of 13.6 *±* 4.5 minutes.

### II-B. Preprocessing

The raw accelerometer data was preprocessed following our previous work [3], which includes cubic spline interpolation to 25 Hz, calibration to 1 g for stationary periods, computation of accelerometer resultant(R) as shown in Eq.1, and high-pass filtering of the resultant using an infinite impulse response (IIR) Butterworth filter with a stopband attenuation of 80 dB at 0.1 Hz and passband ripple of 1 dB starting at 0.5 Hz.

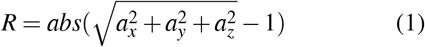

#### II-C. Walk Bout Detection Algorithm

We considered walking bouts of at least 10 seconds meaningful in terms of later analysis and characterization. This is based on the minimal gait duration for trained neurologists evaluating parkinsonism in the clinical setting. Walking bouts are visually recognizable in raw accelerometer data as noisy sinusoidal waves with a quasi-stationary fundamental frequency, which can vary between walking bouts.

Based on these considerations, the resultant (R) was initially segmented into 10-second windows and analyzed through the autocorrelation function (ACF). The ACF was analyzed from a lag between 0.3 and 4 seconds, and the maximum value and location were extracted. A Walking Score (WS) was computed for each window as shown in Eq.2 where *ACF*_*n*_ is the normalized ACF.

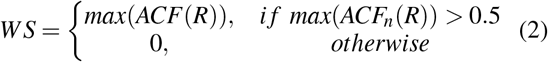

#### II-D. Postprocessing

A post-processing scheme was implemented in two steps. First, a detection threshold was fitted to maximize the F1 score to self-report walking in 7 participants. In this comparison, walking bouts closer than 60 seconds were combined to match the 1-minute resolution of self-reporting. Secondly, the walking bout onset and offset were adjusted to match the data exactly rather than having a 10-second resolution.

This was achieved by first computing the dominant frequency of R matching the walking cadence and subsequently searching for the first significant decrease in the power of that frequency. The walking cadence was extracted for each 10-second window by computing the fast Fourier transform of the ACF and finding the peak frequency. For the first and last 10-second window, the nearest 1-second window (at most 5 seconds away) with a 75% drop in the power of the dominant frequency ± 5Hz. The frequency power was computed using a short-time Fourier transform (STFT) with a window size of 3 seconds, an overlap of 2 seconds, and a Hann window.

### II-E. Walking Bout Analysis

Walking bouts were analyzed to extract the mean and standard deviation of the cadence and arm swing amplitude. The arm swing amplitude was computed by searching R for peaks with a minimum distance of 75% of the reciprocal of the dominant frequency, which was extracted for both positive and negative peaks. The arm swing amplitude for a 10-second window was then defined as the difference in the median between positive and negative peaks of R. Distributions of metrics for walking bouts were compared between participants using the Kruskal-Wallis test to find if there were any variations in the median between participants.

## III. Results

The performance metrics used to evaluate the walking bout detection algorithm were Precision, Recall, and F1 Score calculated as showed in Eq.3,4 and 5 where TP (True Positives), FP (False Positives) and FN (False Negatives).

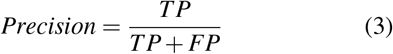

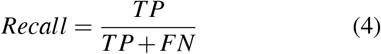

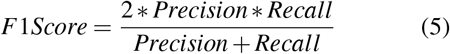

The detection threshold was optimized to be 0.01728 based on the training set, which resulted in an F1 score of 0.66 ±0.08 in the 7 participants. Walk-bout detection results for the individual participants and overall are shown in Table 1.

**Table 1.** Walking bout detector performance on 10-second windows optimized and compared to self-reported walking.

|  | Precision | Recall | F1 |
| --- | --- | --- | --- |
| **Overall** | $0.65 \pm 0.11$ | $0.70 \pm 0.18$ | $0.66 \pm 0.08$ |
| Participant 1 | 0.61 | 0.63 | 0.62 |
| Participant 2 | 0.70 | 0.48 | 0.57 |
| Participant 3 | 0.52 | 0.80 | 0.63 |
| Participant 4 | 0.74 | 0.92 | 0.82 |
| Participant 5 | 0.73 | 0.69 | 0.71 |
| Participant 6 | 0.78 | 0.51 | 0.61 |
| Participant 7 | 0.49 | 0.92 | 0.64 |

After postprocessing using the optimized threshold, walking bouts were compared to a single test recording with all walking bouts carefully annotated with 1-second resolution. This comparison showed an F1 score of 0.84 per area and 0.83 per event considering any overlap as true positives as shown in Table 2.

**Table 2.** Walking bout detector performance compared to the visual annotation of walking.

|  | Precision | Recall | F1 Score |
| --- | --- | --- | --- |
| Per Area | 0.93 | 0.77 | 0.84 |
| Per Event | 0.90 | 0.77 | 0.83 |

As shown in Fig.2, the method has a good agreement with self-reported walking. Although, there are cases of both false negatives and false positives as compared to self-reported walking. In comparison to the visual annotation of walking in the test recording, the annotations match better, of which an example is shown in Fig.3.

**Figure 2.**
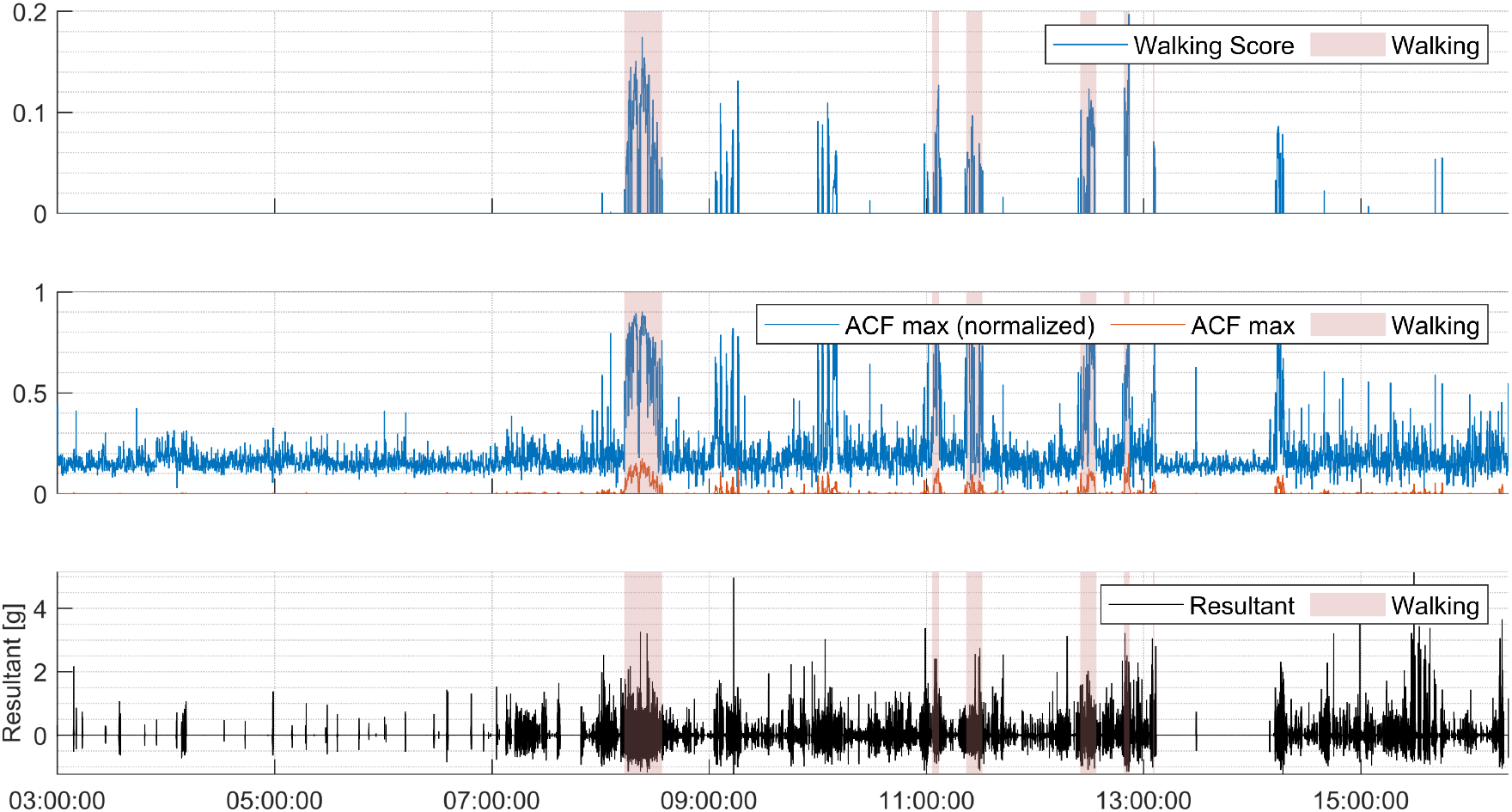
An example tracing of a walking bout in the test recording with precise annotations.

**Figure 3.**
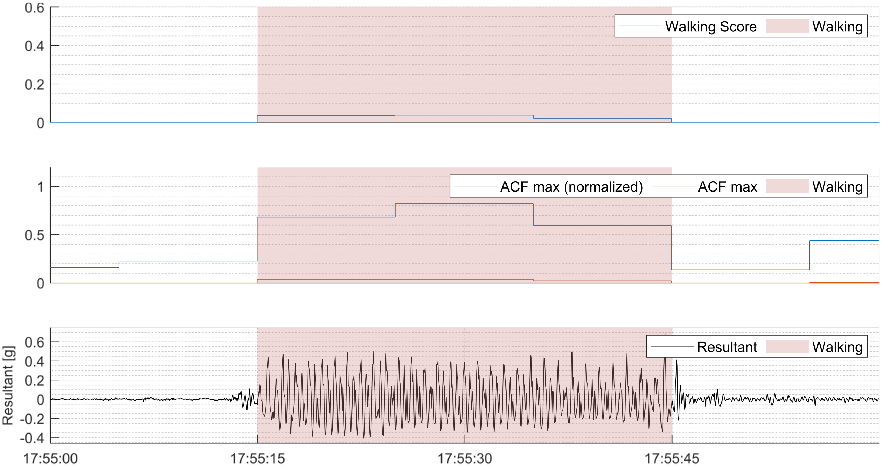
Visual comparison of walking score with self-reported walking activity. The shaded regions indicated the manually marked walking bouts.

The cadence and arm swing amplitude were quantified from detected walking bouts and analyzed for walking bouts with a duration longer than 60 seconds and cadence between 60 − 199 steps/min (Fig.4). The average cadence clearly varies between participants (*p* = 1.19 · 10^*−*10^, however, the standard deviation within walking bouts was stable constant between participants (p = 0.38). Moreover, the mean and within bout standard deviation of arm swing amplitude varied between participants (*p* = 4.83 · 10^*−*18^and *p* = 8.49 · 10^*−*9^, respectively).

**Figure 4.**
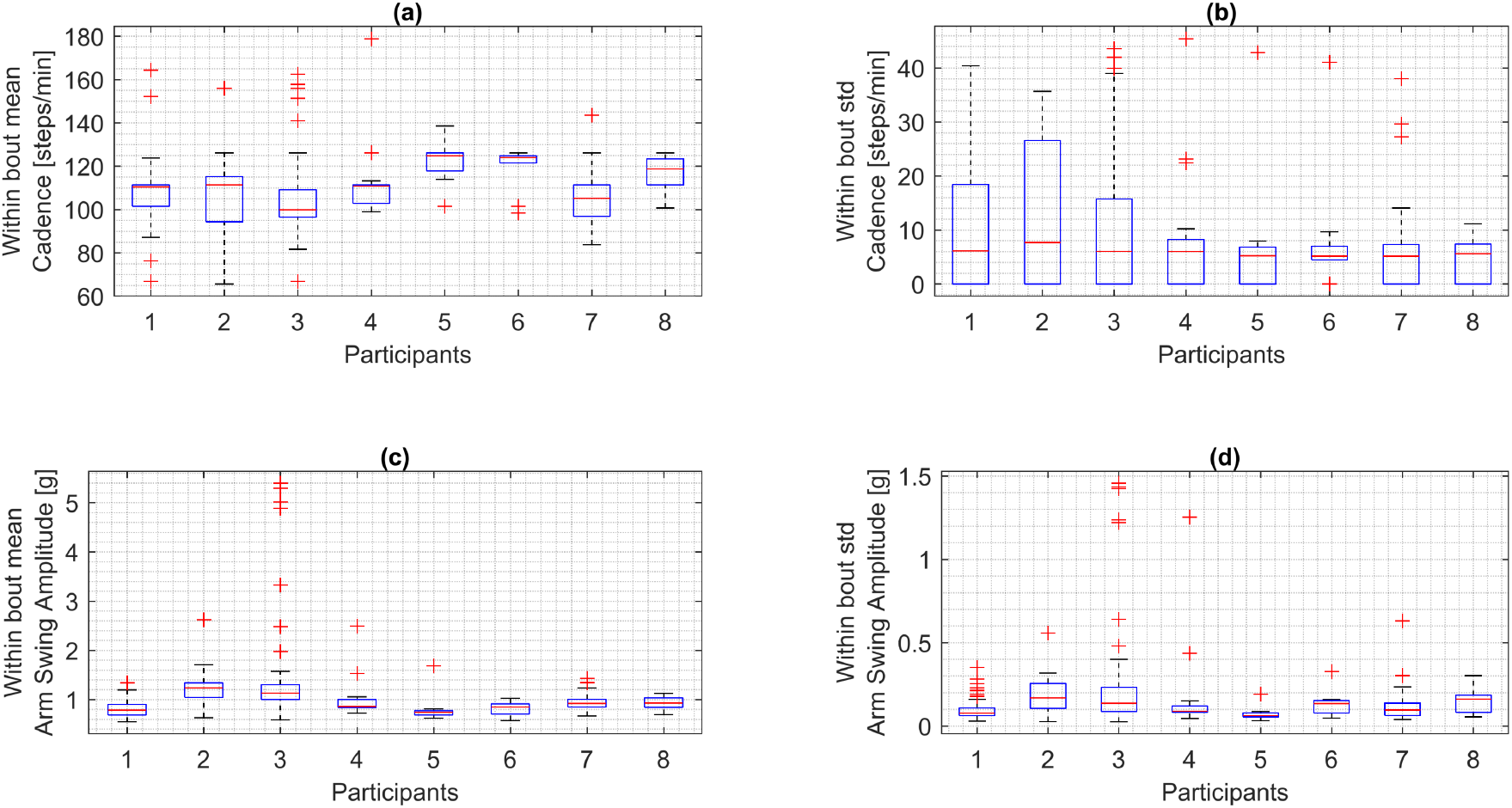
Distribution of walking bout metrics. (a) Average cadence of walking bouts. (b) standard deviation of cadence within walking bouts. (c) Average arm swing amplitude of walking bouts. (d) standard deviation of arm swing amplitude within walking

## IV. Discussion

This study proposes a walking bout detection and characterization algorithm based on the tri-axial accelerometer data obtained passively, in free-living conditions through wrist actigraphy data. The results showed that a gait detection model could achieve good performance, and in particular, high precision in detecting walking bouts. This experiment represents the first step in a systematic analysis of movement characteristics of parkinsonism, which in patients with RBD would signal a high risk of progression to PD.

In comparison to the test recording with high-resolution walking annotations, the detector displayed high precision, which suggests that derived walking characteristics reflect actual walking rather than other movement behaviors. This preliminary pipeline shows the feasibility of automated detection and characterization of gait in free-living conditions using wrist actigraphy. A certain level of false negatives and false positives was to be expected due to imperfect ground truth. False negatives can in part be explained by self-reported walking bouts that in reality included multiple short bouts with breaks in between. False positives can in part be explained by a partial report of walking bouts, particularly shorter ones.

Wrist actigraphy devices are cheap, easily accepted by patients, and allow continuous monitoring during prolonged periods of days to weeks without the need for recharging. Our work supports the feasibility of remote monitoring of gait as a daily behavior expected to show features of parkinsonism, dispensing the need for in-clinic testing or invasive sets of sensors. If proved accurate, this approach may be implemented in large patient cohorts, RBD registries, and large available datasets, such as the UK Biobank.

There are limitations to our study as it was only validated in a small sample without any participants with RBD or PD. Moreover, we did not explicitly test whether and how various external conditions (varying terrain or walking with a phone or bag in one or two hands) could affect the algorithm or derived characteristics. Therefore, future work should include validation in a patient population in various controlled and free-living conditions. Ideally, it should also be compared to a more accurate ground truth, such as video recordings of gait scored by blinded expert clinicians.

Moreover, we showed the feasibility of measuring arm swing amplitude; however other features of gait associated with Parkinsonism should be explored, including U-turns, hesitancy over initiation of gait, and impaired coordination of walking movements.

## V. Conclusions

Detection and characterization of walking bouts is essential for developing a convenient and scalable method for detecting early signs of neurodegenerative diseases, such as PD. Actigraphy has the ability to passively collect data for extended periods of time, non-invasively, and in the patients natural living environment. This paper presents an algorithm that uses single-wrist actigraphy to detect and characterize walking bouts in freeliving conditions, which is the first step toward developing an early detection algorithm for motor changes due to PD. The algorithm can be implemented in larger data cohorts, such as the UK Biobank, which includes actigraphy data for the prediction of health outcomes.

## Data Availability

All data produced in the present study are available upon reasonable request to the authors

## Acknowledgements

This work was supported in part by the Michael J. Fox Foundation grant MJFF-023198 and NIH K25HL151912.

